# THE DISTRIBUTION OF KERATOMETRY READINGS, CORNEAL POWER AND CORNEAL ASTIGMATISM AMONG MALAWIAN YOUNG ADULTS

**DOI:** 10.1101/2023.06.06.23291009

**Authors:** Kenneth Gondwe, Thokozani Mzumara, Joseph Afonne

## Abstract

**Background/objectives:** keratometry and corneal measurements differ among populations. The aim of the study was to assess the distribution of keratometric values and corneal astigmatism and examine the association with age and gender among normal Malawian adults.

**Methods:** This was a cross-sectional study conducted among Mzuzu university students in Malawi. Participants were selected using systematic sampling techniques. K readings were measured using a manual keratometer. Data was entered in SPSS v 26. Chi-square was used to assess association, spearman to assess correction, and an independent t-test to compare the mean.

**Results:** We recruited 98 participants, of whom 59 (60.2%) were male and 39 (39.8%) were female. The Mean age of participants was 27.13 years (SD=5.616). Based on gender, it was 27.97 (SD=5.860) among males and 25.87 (SD=5.038) among females. But an independent t-test showed no significant difference in age according to gender(t(96)= 1.8, p=0.71). On average, the flat and steep K reading was 44.93D (SD=1.49) and 45.40D (SD=1.53) respectively. The mean K was 45.17D (SD=1.47). With regards to gender, the mean K was 45.17D (SD=1.38) and 45.18D (SD=1.62) among males and females respectively, but it was not statistically significant. (t(96)=-0.045, p=0.96). Spearman test showed that the correlation between mean K and age was statistically non-significant mean

**Conclusion:** Corneal astigmatism is relatively tiny among this population group hence surgeons can use techniques to correct and other techniques to achieve greater visual rehabilitation. This study confirms that Environment and genetics play a major role in corneal changes.

## INTRODUCTION

Uncorrected refractive error is the leading cause of visual impairment worldwide. [1] The two major components of refractive error include spherical and astigmatism. [2] Astigmatism can further be classified into lenticular and Corneal Astigmatism (CA).[1] CA is defined as the difference in radius of curvature between two principal meridians., the steep (stronger meridian) and the flat (weaker meridian) meridian. Besides, increased CA is a risk factor for other conditions such as amblyopia and strabismus. Generally, CA is larger among white-skinned people. [1]

Astigmatism greatly contributes to the visual acuity of pseudo-aphakic patients. [3] Accordingly, CA greater than 1 D could prevent optimal vision. [4] Fortunately, toric implantation can correct about O.8 D of CA.[5] Hence, analysis of corneal Astigmatism is relevant for IOL manufacturers. Since the advent of modern cataract surgery techniques enables the correction of astigmatism. [6,7] Knowledge regarding the distribution of CA is crucial for the reduction of post-operative astigmatism during cataract surgery by selecting appropriate methods such as limbal relaxing incisions, opposite clear corneal incisions, and excimer laser refractive procedures. [5] In theatre, different degrees of CA, including magnitude and axis, call for varying approaches to surgical corrections. [6]

Globally, Different populations have varying corneal parameter values. Accordingly, different factors such as genetics, age, gender, and environment have a huge effect on corneal anatomy. [2] For instance, keratometric values are similar among Caucasians and slightly lower in the Far eastern populace. [8] Moreover, values vary according to measuring techniques since various instruments range from manual to automatic devices. [1,8] Nevertheless, to the best of our knowledge there is a paucity of information for the Malawian populace. Hence, this study aims at assessing the distribution of keratometric readings and corneal astigmatism and examine the association with age and gender among normal Malawian adults. The study employed affordable equipment and hence can be useful for optometry and ophthalmology practitioners in the region as a reference.

## METHODS

We conducted a cross-sectional quantitative study among students aged between 18 and 50 years. Based on a sample size determination proposed by Yamane, [9] we recruited 98 participants The study employed stratified systemic sampling where the population was stratified according to faculty. Then, within each faculty participants were recruited considering the nth term. As an exclusion criterion, we included all non-contact lens wearers and excluded participants with a history of ocular surgery and trauma, corneal scar and keratoconus eyes inflammatory eye disease, and other corneal disorders.

### Procedures

We used a Bausch and Lomb keratometer to measure the anterior vertical and horizontal corneal curvature. [10] First, the patient was sat in a standard optometric examination room at room temperature. Prior to collecting demographic information on age and sex, we explained the procedure to the participants. We disinfected the keratometer chin and headrest in between patients. We measured both the right and left eyes. The right eye was measured first, then the left eye. The keratometric parameters included the flat (K1) and steep (K2) and recorded the results in Diopters (D). Each variable was measured 5 times and the average was considered the final value which was recorded on a preform.

### Corneal astigmatism, mean K and axis of astigmatism

We defined CA as the 3 mm steep meridian power minus the 3 mm flat meridian. The axis of the astigmatism was derived from the steep meridian. The CA was considered With the Rule (WTR) when the axis was between 60 and 119 while Against the Rule (ATR) when the axis was between 0 to 29 or 150 to 180 otherwise it was considered oblique. (1) Mean K was computed as the average of the two meridians. Astigmatism was considered a cylinder power of more than 0.5 D. (2)

### Analysis

We entered the data in SPSS version 26. We employed descriptive statistics to describe frequencies and we graphically illustrated the data using graphs, boxplots, and tables. In addition, we execute an independent t-test to compare the mean values of two variables.

Whereas, we utilized one-way ANOVA to compare the mean of variables more than two. Furthermore, we applied the Spearman rank test to explore correlations. Accordingly, we considered the value of p < 0.05 statistically significant.

### Ethics

The study adhered to the declaration of Helsinki. In particular, we obtained ethical approval from the faculty of health sciences research committee. Moreover, we obtained informed consent from each participant and maintained the anonymity of subjects throughout the process. And participant was injured during the study.

## RESULTS

To select the appropriate statistical tools for analysis, a Shapiro-Wilk test was conducted to determine the distribution of the flat and steep K. The test showed K901 (W = 0.989, p < 0.571) and K1802 (W = 0.984, p < 0.268) were normally distributed.

### Demographic and clinical features of study participants

We recruited 98 participants, out of which 59 (60.2%) were male and 39 (39.8%) were female representing a 1.5 to 1 male-to-female ratio. The Mean age of participants was 27.13 years (SD=5.616). Based on gender, it was 27.97 (SD=5.860) among males and 25.87 (SD=5.038) among females, but the independent t-test showed no significant differences in age according to gender(t(96)= 1.8, p=0.71). The K readings in the left and right eye were strongly correlated, hence we considered the results of the right eye only. On average, flat and steep K reading was 44.93D (SD=1.49) and 45.40D (SD=1.53) respectively. The mean K was 45.17D (SD=1.47). With regards to gender, the mean K was 45.17D (SD=1.38) and 45.18D (SD=1.62) among males and females respectively, but it was not statistically significant. (t(96)=-0.045, p=0.96). Spearman test showed that the correlation between mean K and age was statistically non-significant (p=0.245). Figure 1 CA was present in 89 eyes (90.8%). On average the magnitude of CA was 0.7D (SD=0.53). According to gender, the mean CA was 0.6 D (SD=0.51) among males and 0.8 D (SD=0.54) among females. An independent t-test showed that CA was higher among females and the difference was statistically significant (t (96) =- 0.24, p=0.02). About 67 eyes (69.8%) had corneal astigmatism of less than 1 D. (Figure 2). According to age Highest CA (0.76, SD= 0.55) was recorded among the age group 25-29 while the lowest (0.57, SD=0.53) was registered by the 40-44 age group. (Figure 3). Nevertheless, A one-way ANOVA test showed that age group has no impact on CA. F(5,92)=0.34,P=0.812.

**Figure 1.**
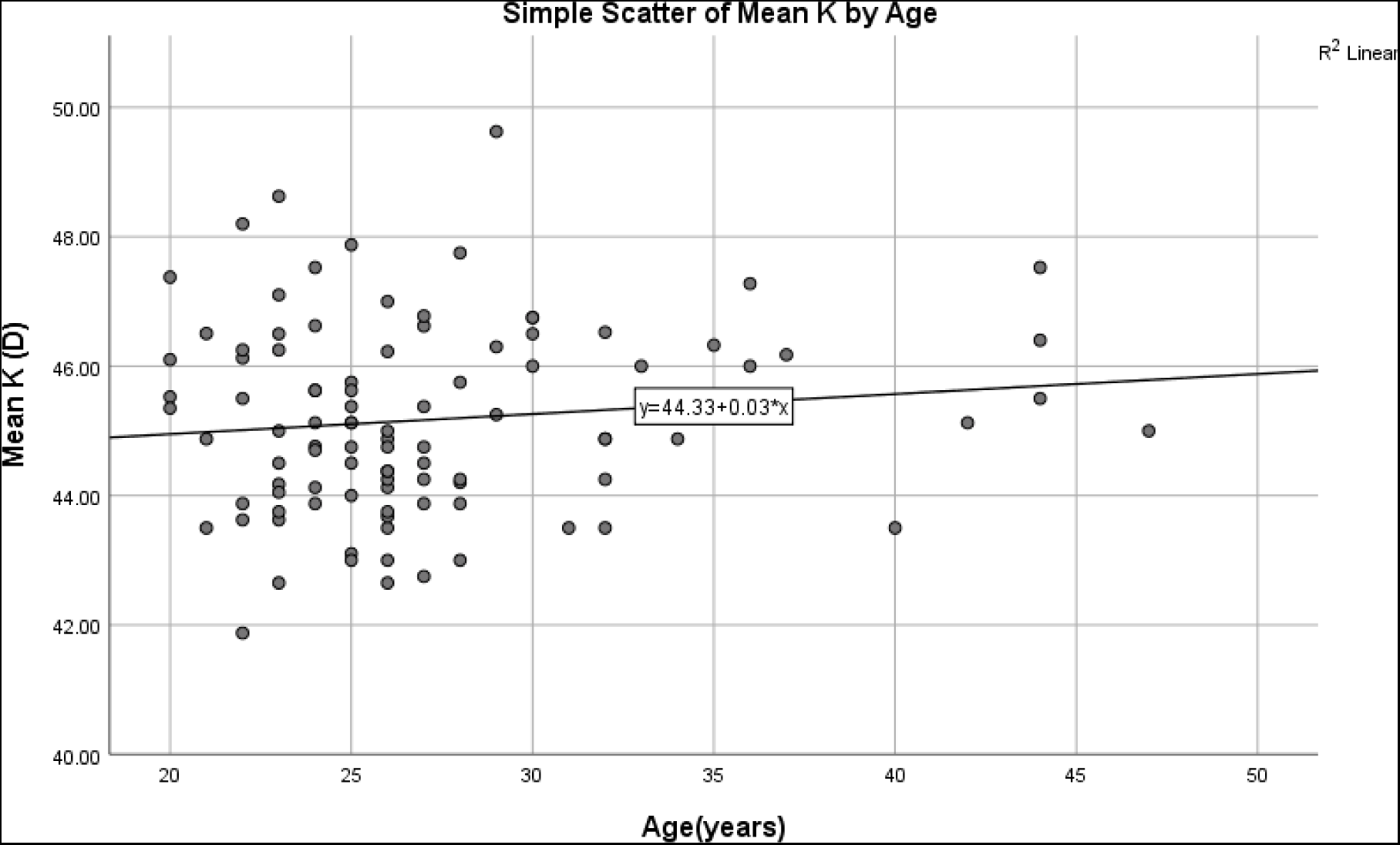
Relationship between mean K and age.

**Figure 2.**
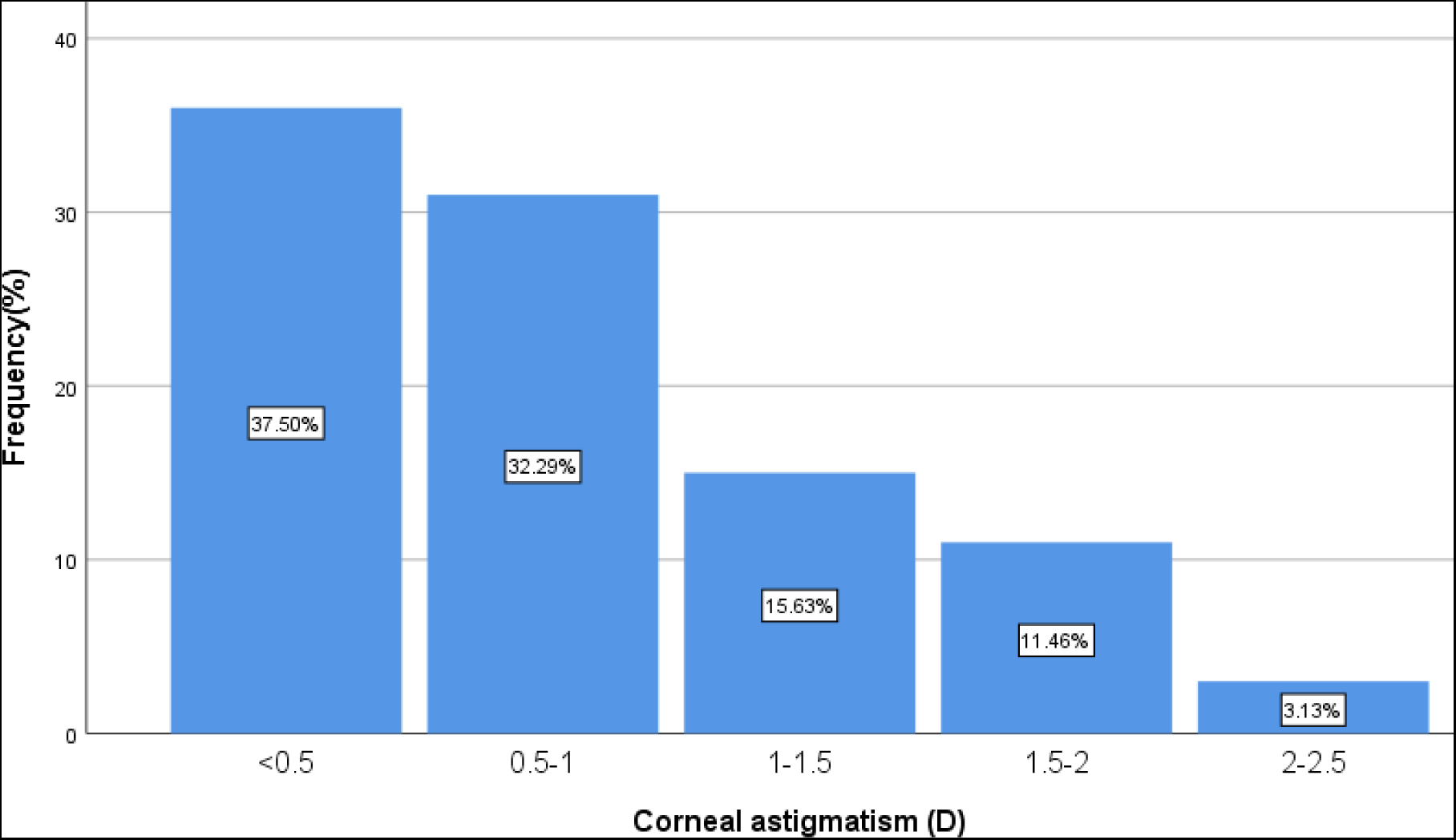
The distribution of corneal astigmatism in 0.50D steps.

**Figure 3:**
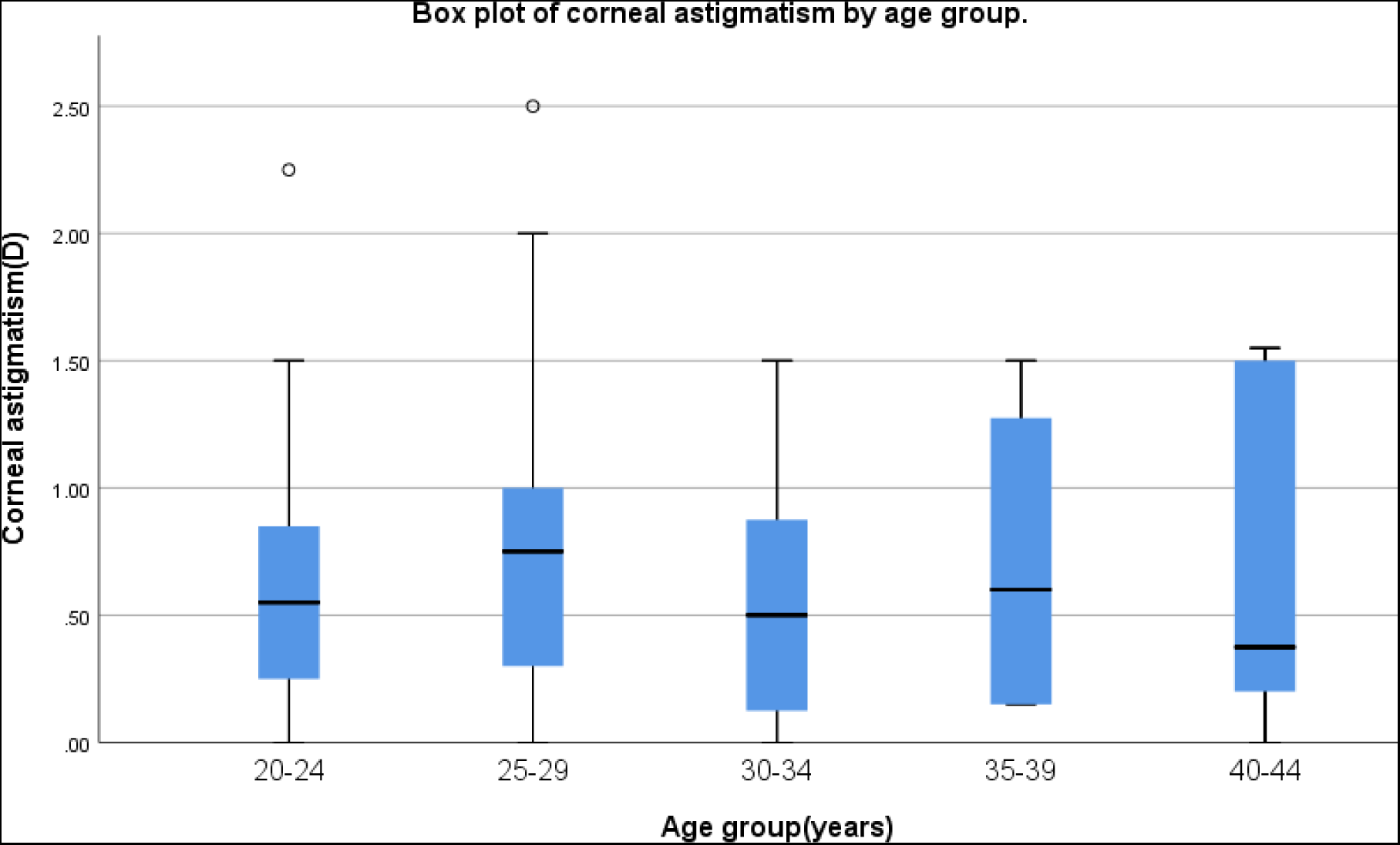
Box plot of corneal astigmatism values (D) in all six age groups; Boxes show the interquartile range. Bold lines in the boxes represent the median (Q2 or 50 % percentile), the upper and lower limits of the box represent the first quartile (Q1 or 25 % percentile) and the third quartile (Q3 or 75 % percentile), and the bars represent the extreme values (maximum and minimum observations). circles represent outliers.

Regarding the type of CA, WTR astigmatism was found in 72 (73.5%) participants, and ATR in 17 (17.3%). Out of the 72 eyes with WTR, 45 (62.5%) were males compared to 27 (37.5%) females. While out of the 17 eyes with ATR, 8 (47.1%) Were males and 9 (52.9%). A chi-square test examined the relationship between gender and type of CA and found that the association was not statistically significant, X2 (1, N = 98) = 1.4, p = 0.243. (Figure 4)

**Figure 4.**
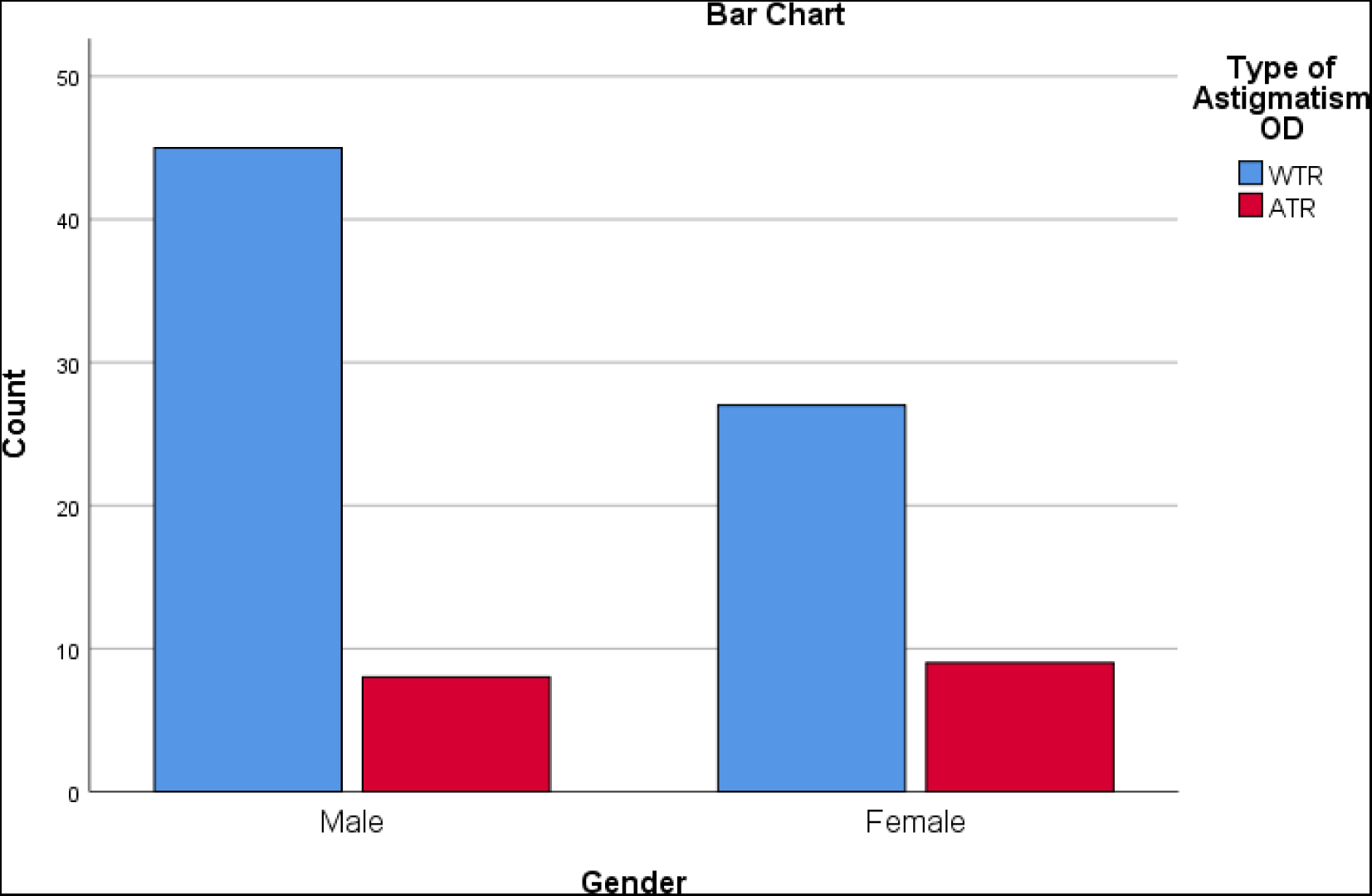
Distribution of WTR and ATR CA between males and females.

## DISCUSSION

Understanding the distribution of corneal values is critical not only for the management of refractive errors but also decision-making in the clinical diagnosis of pathologies such as keratoconus. [11]

In the present study, the mean K was similar to a study in Central China,[5] however Iran[12] found a lower mean K (43.48D), The high mean k in our study could be attributed to higher incidences of myopia in the populace. A recent study [11] found a strong association between myopia and corneal power. Likewise, Thom and colleagues, [13] found that myopia unlike hyperopia is the main cause of reduced vision in Malawi.

With regards to gender, our study found no significant difference between K reading among males and females similar to previous reports. [8,14] Nevertheless, Hashemi [11] found that males have flatter corneas than females. Worldwide there is a general consensus about the gender differences in K readings. The inter-gender difference is explained by anatomical variations in the structure of eyelids and physiological functions mainly due to hormones. [15] The results of our study can be attributed to the ethnicity and genetics of the population group as well as environmental and lifestyle factors [5] Our study included participants of African descent while others included participants of Asian descent. In agreement with previous reports, [16,17] Age had no correlation with K reading in our study. Nevertheless, others [11] found that mean K increases linearly up to the seventh decade of life. The results of our study could be attributed to the narrow age range. Overall, there is strong evidence suggesting an increase in corneal power with age mainly due to physiological changes in corneal biomechanics. [11]

In the current review, the Mean CA was 0.7 D which is lower than in other studies. [8,11] The difference could be attributed to the instrument used. This study used a manual technique while others [8] used an optical refractometer which has relatively more accuracy, precision, and repeatability in biometry measurements. The majority of participants had CA lesser than 0.5 D similar to reports elsewhere. [5] in disagreement, others found a large majority with CA greater than 1 D elsewhere. [18] The low magnitude of CA in our study could be due to ethnicity. Accordingly, Pontikos and colleagues reported that CA was larger among whites and light skin.[1]

According to gender, CA was strongly associated with females more than males similar to previous reports. [1,19] On the contrary, elsewhere, [11,17,21] found no relationship between gender and CA. We cannot explain the findings of our study. With respect to gender, we found that age has no effect on CA similar to previous studies. [11,20] Nevertheless, others have reported that CA decreases according to age. [21] On the other hand, previous studies [3,22] found that CA decreases with age. Astigmatism is associated with younger age. [1] the wide disparity can be explained by the fact that genetics and environmental factors play a major role in corneal structural changes. [11]

Our study found that the majority of eyes had WTR astigmatism similar to the previous study. [23] However, other authors [5,11] found that a majority of eyes had ATR astigmatism. We attribute the variation to the age composition of the two studies. Apparently, we recruited a relatively younger group (mean age= 27 years) compared to Ferreira and colleagues. [8] (mean age=60 years). Globally, It is generally accepted that WTR Astigmatism decreases while ATR increases with age due to anatomical changes in the eyelids and cornea. [11] Our study did not find a relationship between age and orientation of CA in contrast to other authors who suggest that ATR astigmatism increased with age while WTR astigmatism decreased with age, [8,11,13,23,24] Our review found no association between the orientation of CA and gender similar to studies elsewhere. [3] In contrast, others [11] reported that WTR is associated with the female gender whereas, ATR is greater in male eyes. [12]

### Limitations

Our study is not without limitations. First, we employed a narrow age range which could mask the effect of age on the parameters. In addition, the current review did not take into account the posterior corneal surface measurements underestimating total corneal astigmatism. We propose a large-scale study probably at a national level to further investigate this phenomenon. Furthermore, our study results could be overestimated since they used manual measurements of k readings. Our study did not assess the correlation between refractive status and K readings.

## CONCLUSION

The study provides normal ocular biometric measurements with regard to K readings and anterior corneal astigmatism and the effects of age. The study found that Age and gender are not related to keratometry readings. CA was found to be associated with the female gender but not age. The majority have CA < 1.00D, which can be corrected by low-cost procedures like steep axis phaco, limbal relaxing incisions, and opposite clear corneal incisions, especially in developing countries like Malawi. In addition, less percentage of candidates would require expensive toric IOLs. The Majority of eyes are WTR astigmatism. Our study suggests that ocular biometry should be considered as a pre-operative routine procedure to help improve visual outcomes and reduce the dependency on glasses for this underprivileged population.

## Data Availability

All data produced in the present study are available upon reasonable request to the authors

